# The Safety and Efficacy of Cangrelor in Endovascular Thrombectomy Compared with Glycoprotein IIb/IIIa Inhibitors

**DOI:** 10.1101/2024.06.14.24308962

**Authors:** Alex Devarajan, Shouri Gottiparthi, Michael T. Caton, Aya Ouf, Katty Wu, Daryl Goldman, Nicole Davis, Nadine Musallam, Jack Y. Zhang, Naina Rao, Neha Dangayach, Connor Davy, Michael Fara, Shahram Majidi, Thomas Oxley, Christopher Kellner, Tomoyoshi Shigematsu, Reade DeLeacy, J Mocco, Johanna T. Fifi, Hazem Shoirah

## Abstract

**Background:** Cangrelor, an intravenous P2Y12-receptor inhibitor, is a reversible and short-acting antithrombotic medication non-inferior to irreversible Glycoprotein IIb-IIIa inhibitors (GPI) like eptifibatide. However, there is insufficient data to compare the medications in endovascular thrombectomy (EVT) cases requiring emergent platelet inhibition. This study reviews our experience with cangrelor in EVT and compares its safety and efficacy against GPIs.

**Methods:** A large healthcare system retrospective review identified all patients who received cangrelor or eptifibatide intraoperatively during EVT from December 2018 to March 2023 for a cohort study. Clinical data was reviewed. Functional status was defined by the modified Rankin Scale (mRS) and National Institutes of Health Stroke Scale (NIHSS) at multiple time points. Binary variables were tested with Pearson χ2 tests or Fisher’s exact tests. Continuous variables were tested with two-tailed t-tests or Wilcoxon tests.

**Results:** Of 1,010 EVT patients, 36 cangrelor and 104 eptifibatide patients were selected. There were no significant differences in baseline functional status or presentations. Cangrelor was most frequently administered for stenting tandem occlusions (n=16, 44.4%) and successful reperfusion occurred in 93.3% of patients (n=30). On multivariate analysis, cangrelor usage was associated with decreased odds of hemorrhagic conversion (adjusted odds ratio (AOR) 0.76, p=0.004) and symptomatic hemorrhage (AOR 0.86, p=0.021). There were no significant differences in thrombotic re-occlusion. Cangrelor was associated with lower 24-hour NIHSS (7.0 vs. 12.0, p=0.013) and discharge NIHSS scores (3.0 vs 9.0, p=0.009). There were no significant differences in in-hospital mortality or length of stay. Cangrelor was associated with improved odds of favorable outcome, defined as mRS 0-2, at discharge (AOR 2.69, p=0.001) and on 90-day follow-up (AOR 2.23, p=0.031).

**Conclusion:** Cangrelor was associated with a decreased risk of hemorrhagic conversion and may lead to favorable functional outcomes for patients during hospitalization when compared to GPIs. Future prospective studies are warranted to investigate its use in EVT.

**Previous Presentations:** This abstract was previous presented as a podium presentation at the Society of Neurointerventional Surgery’s 20^th^ Annual Meeting in San Diego, CA from July 31^st^ - August 4^th^, 2023.

## Introduction

Endovascular thrombectomy (EVT) is considered to be the standard of care for patients suffering acute ischemic stroke due to large vessel occlusion.^1–3^ Within the last decade, the indications for EVT have continued to expand. While the safety and efficacy of EVT are well established, certain conditions may necessitate immediate platelet inhibition, such as patients presenting with tandem occlusions, concomitant large vessel occlusion with extracranial stenosis or occlusion, or patients with acutely symptomatic intracranial atherosclerotic disease (ICAD).^4^ Tandem lesions constitute approximately 10% of all ischemic stroke presentations and respond poorly to traditional intravenous thrombolysis.^5^ Furthermore, patients with ICAD present with increased risk for morbidity and mortality during stroke recovery.^6–8^ It is not feasible to rely on oral administration of antiplatelet agents, given the emergent nature of those procedures, the patient’s inability to swallow and the prolonged time to action of enterally administered agents. In such cases, intra-arterial administration of platelet GPIIb-GPIIIa receptor inhibitors (GPI) such as eptifibatide and tirofiban is frequently used to prevent re-occlusion by limiting fibrinogen binding.^9^ Though the administration of intraprocedural GPIs provide therapeutic benefit, a feared complication of utilizing GPIs is the substantially increased risk of hemorrhagic conversion due to their partial thrombolytic properties and long half-life.^10^ The irreversible mechanism of action of GPIs further complicates their use, as it limits the effectiveness of platelet transfusions in case of occurrence of any hemorrhagic complications. The risk of symptomatic intracranial hemorrhage (ICH) in stroke patients undergoing interventions with the use of GPIs is reported to be as high as 18%, with a mortality rate of 44%.^11^

Cangrelor, an intravenous P2Y12-receptor inhibitor, is a reversible and short-acting antithrombotic medication approved by the United States Food and Drug Administration for the prevention of thrombotic complications in cardiological interventions.^12–15^ In the cardiac literature, pooled analyses have demonstrated that cangrelor is non-inferior to GPIs in composite mortality and stent thrombosis while demonstrating a lower risk of major or minor bleeding.^10^ Multiple preliminary studies have described the safety and efficacy of cangrelor in neurointervention for a variety of indications, but no study to date has compared its performance against the well-established GPIs.^16,17^ Cangrelor may provide a safe and efficacious alternative to GPIs in the emergent stroke setting.^18–21^ This study reviews our institution’s experience with cangrelor in EVT and compares its safety and efficacy against GPIs.

## Methods

### Cohort Identification and Selection

A large healthcare system retrospective review of a prospectively maintained database identified all patients who received cangrelor or eptifibatide intraoperatively while undergoing EVT for management of acute stroke from December 2018 to March 2023. This retrospective study was approved by the senior author’s Institutional Review Board (IRB) under STUDY# IRB-19-02829. Consent was waived for all patients. This study follows the Strengthening the Reporting of Observational Studies in Epidemiology (STROBE) guidelines for retrospective cohort studies.

### Data Collection and Outcomes

Baseline clinical and demographic data was reviewed, including age, sex, prior antiplatelet/anticoagulant use, diabetes, history of a prior stroke, coronary artery disease (CAD), hypertension, hyperlipidemia, obesity, congestive heart failure (CHF), and smoking status. Clinical characteristics of the initial stroke presentation were reviewed, including administration of IV thrombolysis, stroke location, and whether a tandem occlusion was present. The proceduralist performing the EVT was recorded and broadly stratified into three categories based on number of years of experience as a fellowship-trained attending. When appropriate, an Alberta stroke program early CT score (ASPECTS) score was calculated using pre-procedure non-contrast CT (Computerized Tomography) imaging of the head.^22^ Stroke severity was assessed using the 42-point National Institutes of Health Stroke Scale (NIHSS) at initial presentation to the hospital, 24 hours after EVT, and on discharge. Functional status was assessed at pre-stroke baseline, at discharge, and on 90-day follow-up using the 6-point modified Rankin Scale (mRS). Procedural details including indication for antithrombotic use and choice of oral transition agent were recorded. Indications for the use of cangrelor or eptifibatide were broadly categorized into three groups: placement of an intracranial stent, placement of an extracranial stent, or use for distal recanalization. Hospital course including length of stay in the ICU, in-hospital mortality, total hospital stay, and discharge disposition were reviewed.

Post-procedural outcomes were also assessed. The presence of hemorrhagic transformation was assessed by a board-certified neuroradiologist (MTC) blinded to the patient’s treatment and graded according to the European Cooperative Acute Stroke Study (ECASS) II categories.^23,24^ Hemorrhagic transformation was defined as a symptomatic hemorrhage if it was graded as a hemorrhage per ECASS criteria and was accompanied with a gain in NIHSS of at least 4 points.^24,25^ Vessel and stent patency were assessed for thrombotic complications post-procedure with one of four vascular imaging modalities obtained after EVT: CT angiography, MR angiography, carotid ultrasound, or subsequent cerebral angiogram. The median time to post-procedure vascular imaging, ICU length of stay, and total hospital length of stay with respective interquartile ranges were calculated to minimize skew. mRS scores at discharge and on 90-day follow-up were dichotomized into favorable and unfavorable outcomes, defined by mRS 0-2 and mRS greater than 3 respectively.

### Cangrelor Administration and Bridging

Intravenous cangrelor was administered intra-procedurally according to the manufacturer instructions.^26^ An initial bolus of 30 mcg/kg was administered, followed by a steady infusion rate of 4 mcg/kg/min.

Two medications were used as transition agents when bridging from IV cangrelor. Ticagrelor is started with a 180 mg oral loading dose 1-2 hours prior to discontinuing cangrelor, followed by a 90 mg dose every 12 hours. Clopidogrel was started with a 600 mg dose loading dose, given concomitant with discontinuation of cangrelor, followed by a 75 mg dose administered daily. For all patients, aspirin was administered post-procedurally with a rectal 600 mg loading dose, followed by 325 mg daily, for patients who are transitioned to clopidogrel, or 81 mg daily, for patients transitioned to ticagrelor.

### Statistical Analysis

All statistical analyses were performed in R version 4.2.3. Univariate analysis was initially conducted of the cohort. Categorical variables were tested with Pearson χ2 tests or Fisher’s exact tests if any group had less than 5 data points. Continuous variables were tested with two-tailed t-tests or Wilcoxon rank-sum tests depending on the distribution of the data. Following univariate analysis of the cohort, the study outcomes were analyzed by multivariate regression after controlling for age, gender, comorbidities, antiplatelet/anticoagulant use, occlusion location, the presence of a tandem occlusion, the placement of any stent, IV thrombolysis administration, premorbid mRS, NIHSS scores at initial presentation, and ASPECTS scores. Adjusted odds ratios (AOR) or β-coefficients with 95% confidence intervals were generated for logistic and linear regressions, respectively. Variable collinearity was assessed by testing for variance inflation factors (VIF).

The corresponding (AD) and senior author (HS) had full access to all data collected and analyzed in the study, and take responsibility for its analysis and integrity.

## Results

Among 1,010 patients who underwent EVT during the timeframe of this study, 36 patients who received cangrelor and 104 patients who received eptifibatide intraoperatively were identified and included.

### Patient Demographics and Medical History

The mean age of the cangrelor- and eptifibatide-receiving groups was 65.0 and 65.1 respectively (p=0.988). No statistically significant differences in medical histories were observed between groups, including prior antiplatelet use (27.8% vs 28.8%, p=0.904), prior anticoagulant use (2.8% vs 3.8%, p=0.768), and prior stroke (11.1% vs 12.5%, p=1.000). Baseline functional status as defined by mRS was also not found to be significantly different between groups, with both groups having a median mRS of 0 and interquartile range (IQR) of 0-1 (p=0.806). All patient demographic and medical history information is further outlined in **Table 1**.

**Table 1:** Patient Demographics, Comorbidities, and Medical History.

| **Table 1: Patient Demographics, Comorbidities, and Medical History** |  |  |  |
| --- | --- | --- | --- |
|  | Cangrelor (n=36) | Eptifibatide (n=104) | p-value |
| Age (Mean ± SD) | 65.0 ± 15.0 | 65.1 ± 13.1 | 0.988 |
| Male sex | 24<br>(66.7%) | 71 (68.2%) | 1.000 |
| Premorbid mRS (IQR) | 0 (0, 1) | 0 (0, 1) | 0.806 |
| Prior antiplatelet use | 10<br>(27.8%) | 30 (28.8%) | 0.904 |
| Prior anticoagulant use | 1 (2.8%) | 4 (3.8%) | 0.768 |
| Diabetes | 13 (36.1%) | 41 (39.4%) | 0.725 |
| Prior stroke | 4 (11.1%) | 13 (12.5%) | 1.000 |
| CAD | 7 (19.4%) | 19 (18.3%) | 0.876 |
| Hypertension | 29 (80.6%) | 70 (67.3%) | 0.132 |
| Hyperlipidemia | 15 (41.7%) | 46 (44.2%) | 0.789 |
| Obesity | 14 (38.9%) | 42 (40.4%) | 0.875 |
| CHF | 6 (16.7%) | 15 (14.4%) | 0.745 |
| Smoking | 18 (50%) | 53 (51.0%) | 0.921 |

### Initial Stroke Presentation and Antithrombotic Usage

Indication for administration of antithrombotic agent was significantly different between cangrelor and eptifibatide groups (**p<0**.**001**). Out of the 36 cases in which cangrelor was used, it was used for intracranial stents 13 times (36.1%), extracranial stents 21 times (58.3%), and as a recanalization aid two times (5.6%). Out of the 104 cases in which eptifibatide was used, it was used for intracranial stents 36 times (34.6%), extracranial stents 22 times (22.1%), and as a recanalization aid 46 times (44.2%). No significant differences were observed between cangrelor- and eptifibatide-receiving groups in initial stroke presentation. Median NIHSS at presentation was 12.5 in the cangrelor group (IQR 9.5, 17.0) and 14.0 in the eptifibatide group (IQR 7.0, 19.0; p=0.768). In the cangrelor group, 91.7% of patients had anterior circulation occlusions and 44.4% had tandem occlusions, compared to 79.8% and 28.8% respectively in the eptifibatide group (p=0.170, 0.086). There was no difference in ASPECTS between groups, with initial median ASPECTS score being 9.0 with an IQR of 8.0-10.0 in both groups (p=0.647). There was also no significant difference in administration of IV thrombolysis prior to EVT, with 30.6% of patients in the cangrelor group receiving IV thrombolysis compared to 26.0% in the eptifibatide group (p=0.596). Proceduralist experience also did not affect the choice to utilize cangrelor or eptifibatide (p=0.442).

The transition agent used after discontinuation of the antithrombotic agent differed significantly between groups **(p<0**.**001)**. No transition agent was used after cangrelor only 11.1% of the time compared to 41.3% after eptifibatide. Ticagrelor, rather than clopidogrel, was more frequently used after cangrelor (47.2% vs 2.9%), while clopidogrel was more frequently used after eptifibatide (41.3% vs 11.1%). All collected data regarding initial stroke presentation and management is outlined in **Table 2**.

**Table 2:** Initial Stroke Presentation and Management.

| **Table 2: Initial Stroke Presentation and Management** |  |  |  |
| --- | --- | --- | --- |
|  | Cangrelor (n=36) | Eptifibatide (n=104) | p-value |
| NIHSS at presentation (Median (IQR)) | 12.5 (9.5, 17.0) | 14.0 (7.0, 19.0) | 0.768 |
| Baseline ASPECTS (Median (IQR)) | 9.0 (8.0, 10.0) | 9.0 (8.0, 10.0) | 0.647 |
| IV thrombolysis used | 11 (30.6%) | 27 (26.0%) | 0.596 |
| Anterior circulation occlusion | 33 (91.7%) | 83 (79.8%) | 0.170 |
| Tandem occlusion | 16 (44.4%) | 30 (28.8%) | 0.086 |
| Proceduralist experience |  |  | 0.442 |
| Less than 5 years | 1 (2.8%) | 1 (1.0%) |  |
| 5 to 15 years | 26 (72.2%) | 67 (64.4%) |  |
| Greater than 15 years | 9 (25.0%) | 36 (34.6%) |  |
| Indication for administration |  |  | **<0.001** |
| Intracranial stent | 13 (36.1%) | 36 (34.6%) |  |
| Extracranial stent | 21 (58.3%) | 22 (21.1%) |  |
| Recanalization aid | 2 (5.6%) | 46 (44.2%) |  |
| Transition agents used |  |  | **<0.001** |
| Ticagrelor | 17 (47.2%) | 3 (2.9%) |  |
| Clopidogrel | 15 (41.7%) | 58 (55.8%) |  |
| No transition agent | 4 (11.1%) | 43 (41.3%) |  |

### Univariate Analysis of Outcomes

The rate of overall hemorrhagic transformation after EVT was significantly lower in the cangrelor-receiving group compared to the eptifibatide-receiving group (19.4% vs 42.3%, **p=0**.**026**). Two patients in the cangrelor group had a PH-2 transformation (5.6%), and 8 patients in the eptifibatide group had a PH-2 transformation (7.7%). The rates of symptomatic hemorrhagic were not significantly different between groups (8.3% vs 17.3%, p=0.280). Vascular imaging was not available for 6 cangrelor patients and 34 eptifibatide patients to assess thrombotic re-occlusion. No significant difference between groups in rates of thrombotic re-occlusion after EVT (6.7% vs 20.0%, p=0.126). The cangrelor group was found to have lower median NIHSS at both 24 hours after EVT (7 vs 12, **p=0**.**013**) and at discharge (3 vs 9, **p=0**.**004**). No significant differences were observed between groups in in-hospital mortality 8.3% vs 13.5%, p=0.560), length of stay in the ICU (6.0 days vs 6.2 days, p=0.852), and total length of hospitalization (10.0 days vs 11.0 days, p=.610). More patients in the cangrelor group were functionally independent, mRS 0-2, at both discharge (44.4% vs 25.0%, **p=0**.**028**) and at 90-day follow up (58.3% vs 30.2%, **p=0**.**003**). 10 patients in the eptifibatide group were lost to follow-up and a 90-day mRS could not be obtained. There was no difference in in-hospital mortality (8.3% vs 13.5%, p=0.560). All univariate analyses are outlined in **Table 3**.

**Table 3:**
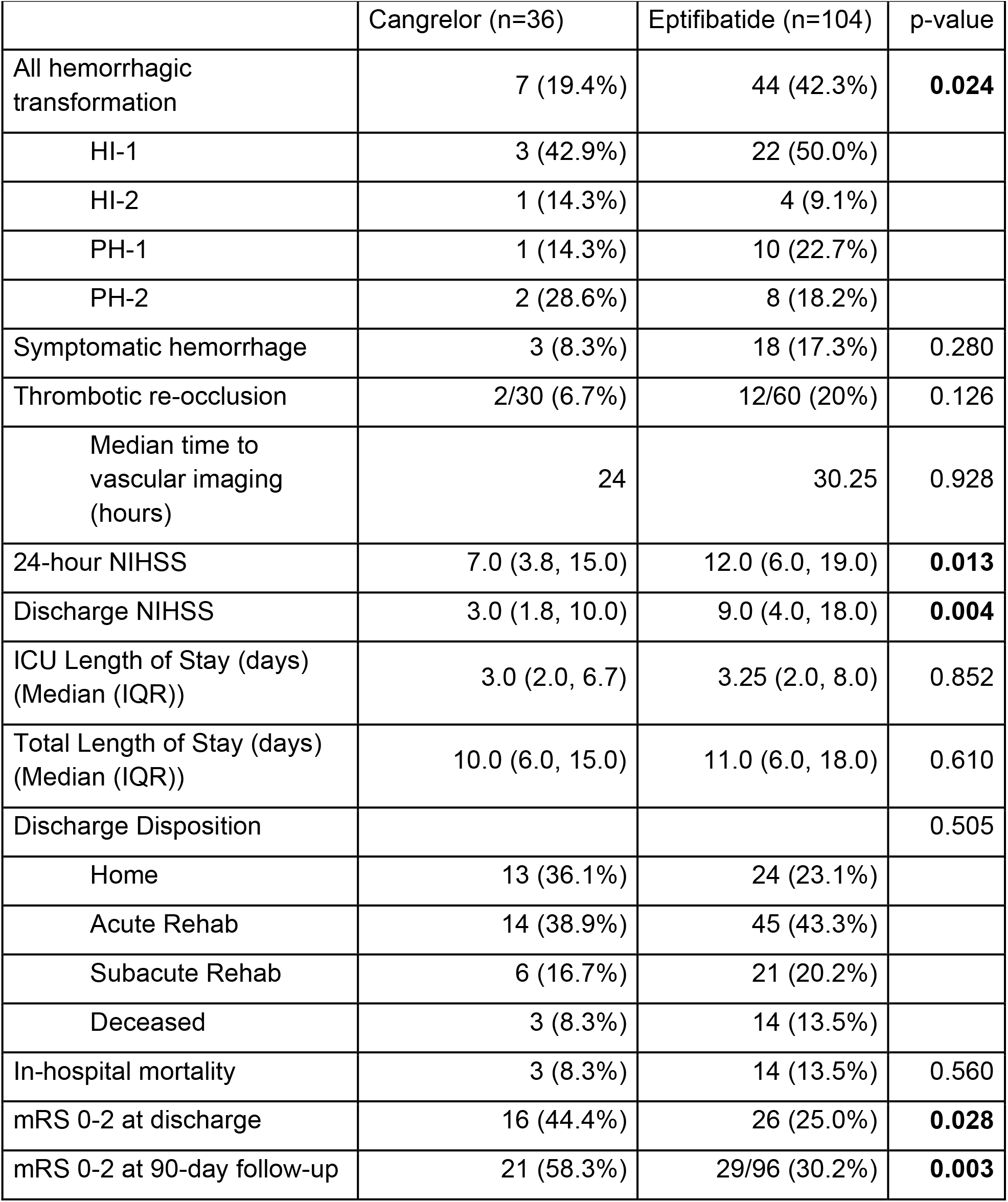
Univariate Analysis of Outcomes.

| **Table 3: Univariate Analysis of Outcomes** |  |  |  |
| --- | --- | --- | --- |
|  | Cangrelor (n=36) | Eptifibatide (n=104) | p-value |
| All hemorrhagic transformation | 7 (19.4%) | 44 (42.3%) | **0.024** |
| HI-1 | 3 (42.9%) | 22 (50.0%) |  |
| HI-2 | 1 (14.3%) | 4 (9.1%) |  |
| PH-1 | 1 (14.3%) | 10 (22.7%) |  |
| PH-2 | 2 (28.6%) | 8 (18.2%) |  |
| Symptomatic hemorrhage | 3 (8.3%) | 18 (17.3%) | 0.280 |
| Thrombotic re-occlusion | 2/30 (6.7%) | 12/60 (20%) | 0.126 |
| Median time to vascular imaging (hours) | 24 | 30.25 | 0.928 |
| 24-hour NIHSS | 7.0 (3.8, 15.0) | 12.0 (6.0, 19.0) | **0.013** |
| Discharge NIHSS | 3.0 (1.8, 10.0) | 9.0 (4.0, 18.0) | **0.004** |
| ICU Length of Stay (days) (Median (IQR)) | 3.0 (2.0, 6.7) | 3.25 (2.0, 8.0) | 0.852 |
| Total Length of Stay (days) (Median (IQR)) | 10.0 (6.0, 15.0) | 11.0 (6.0, 18.0) | 0.610 |
| Discharge Disposition |  |  | 0.505 |
| Home | 13 (36.1%) | 24 (23.1%) |  |
| Acute Rehab | 14 (38.9%) | 45 (43.3%) |  |
| Subacute Rehab | 6 (16.7%) | 21 (20.2%) |  |
| Deceased | 3 (8.3%) | 14 (13.5%) |  |
| In-hospital mortality | 3 (8.3%) | 14 (13.5%) | 0.560 |
| mRS 0-2 at discharge | 16 (44.4%) | 26 (25.0%) | **0.028** |
| mRS 0-2 at 90-day follow-up | 21 (58.3%) | 29/96 (30.2%) | **0.003** |

### Multivariate Analysis of Outcomes

On multivariate analysis, use of intraoperative cangrelor rather than intraoperative eptifibatide was now associated with lower odds of symptomatic hemorrhage after EVT (AOR 0.86, CI 0.75-0.97, **p=0**.**021**). Cangrelor use continued to be associated with lower odds of overall hemorrhagic transformation after EVT (AOR 0.76, CI 0.63-0.91, **p=0**.**004**). Cangrelor use was not observed to significantly influence the odds of thrombotic reocclusion after EVT (AOR 1.13, CI 0.96-1.34, p=0.146). Cangrelor use was associated with lower NIHSS 24 hours after EVT (β 0.24, CI 0.13-1.10, **p=0**.**013**) and at discharge (β 0.17, CI 0.10-0.67, **p=0**.**009**). Cangrelor use was not associated with in-hospital mortality (AOR 0.93, CI 0.82-1.05, p=0.229), length of stay in the ICU (β 0.05, CI -2.45-2.55, p=0.967), or total length of stay (β -0.90, CI -9.92-8.13, p=0.844). Cangrelor use was associated with improved odds of having functional status classified as mRS 0-2 at discharge (AOR 2.69, CI 1.48-4.87, **p=0**.**001**) and 90-days after discharge (2.23, CI 1.04-5.10, **p=0**.**031**). All multivariate analyses are outlined in **Table 4**.

**Table 4:**
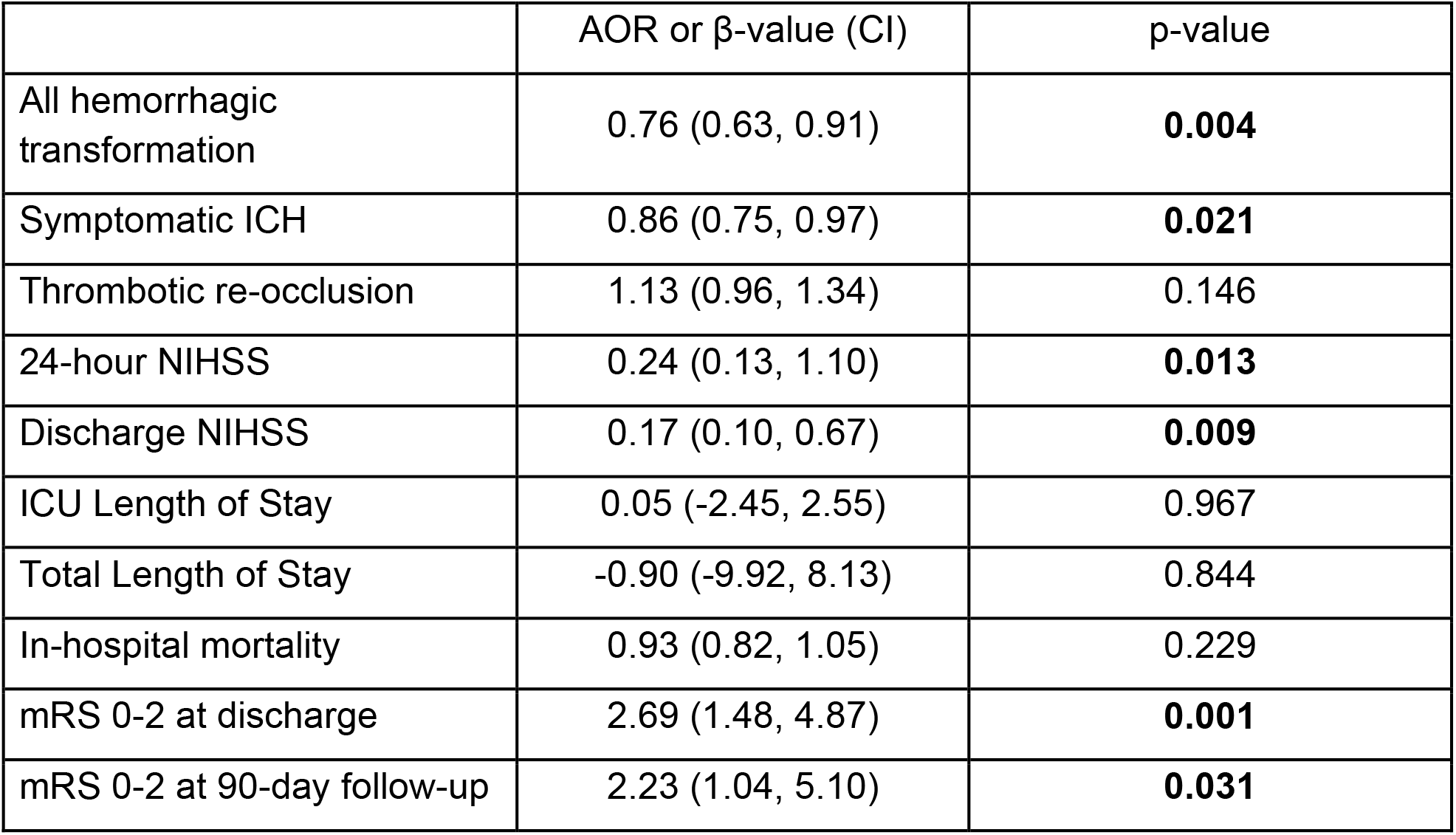
Multivariate Analyses of Outcomes.

| | AOR or β-value (CI) | p-value |
| --- | --- | --- |
| All hemorrhagic transformation | 0.76 (0.63, 0.91) | **0.004** |
| Symptomatic ICH | 0.86 (0.75, 0.97) | **0.021** |
| Thrombotic re-occlusion | 1.13 (0.96, 1.34) | 0.146 |
| 24-hour NIHSS | 0.24 (0.13, 1.10) | **0.013** |
| Discharge NIHSS | 0.17 (0.10, 0.67) | **0.009** |
| ICU Length of Stay | 0.05 (-2.45, 2.55) | 0.967 |
| Total Length of Stay | -0.90 (-9.92, 8.13) | 0.844 |
| In-hospital mortality | 0.93 (0.82, 1.05) | 0.229 |
| mRS 0-2 at discharge | 2.69 (1.48, 4.87) | **0.001** |
| mRS 0-2 at 90-day follow-up | 2.23 (1.04, 5.10) | **0.031** |

## Discussion

This study demonstrates the utility of cangrelor as an antiplatelet option during EVT, with comparable efficacy to GPIs and an attractive safety profile. After controlling for multiple baseline and procedural confounding variables, cangrelor use led to decreased rates of hemorrhagic transformation and symptomatic hemorrhage.

Cangrelor use was associated with improved NIHSS and mRS scores at multiple time points during hospitalization and on long-term follow-up without any increased risk of thrombotic complications. Though further prospective studies are needed, these results suggest that cangrelor may be a safer option for rapid and reversible antiplatelet therapy in the emergent stroke setting.

Cangrelor offers many advantages as an antiplatelet medication. Its lack of thrombolytic properties, seen in GPIs, may reduce the risk of hemorrhagic complications, as demonstrated in this study. Given the rapid onset of cangrelor, its infusion can be precisely timed during the procedure to start prior to stent placement.^27^ Additionally, its reversibility and rapid elimination time ameliorates the need for complex interventions or platelet transfusions in case of hemorrhagic complications, unlike GPIs.^14,28^ Cangrelor activity can be monitored using P2Y12 assays to assess platelet inhibition, unlike GPIs, which aids in safe transitions to oral therapy while maintaining therapeutic inhibition.

Other case series have supported the safety and efficacy profile of cangrelor in neurointervention, citing a low rate of hemorrhagic complications with low thrombotic complications.^18,20,27,29–31^ This safety and efficacy profile extends beyond the stroke setting to other neurointerventional procedures, such as the placement of flow-diverting stents or for stent-assisted coiling of aneurysms.^17,19,32,33^ Some of the limitations of those studies include the heterogenous indications for cangrelor use, lack of clarity on status of device patency post-operatively, as well as insufficient reporting of dosing strategies.

In our system, we use the standard dosing regimen of cangrelor administration with a 30 mcg/kg bolus, followed by an infusion of 4 mcg/kg/min. Other publications have reported on using reduced doses of cangrelor. For example, one case series of 12 patients described the use of half-dose intraprocedural cangrelor in the emergent stroke setting with no resulting symptomatic hemorrhagic or thrombotic complications, even with accompanying intracranial stent placement.^17^ However, two other case series described reduced-dosing strategies, either by PRU-based (platelet reactivity unit) dose titration or low-dose boluses, with subsequent thrombotic complications in patients requiring re-thrombectomy.^30,31^ At our own institution, PRU levels were initially collected to monitor therapeutic inhibition. However, this collection was paused as we anecdotally found that the PRU levels did not impact our management decisions. This is a topic that merits further investigation in future studies.

The choice of transition agent plays a significant role in the outcomes of patients who received cangrelor. Cangrelor competes at the P2Y12 receptor with clopidogrel, which may prevent activation of clopidogrel metabolites and reduce the effectiveness of clopidogrel as an antiplatelet agent during cangrelor washout.^36,37^ For that reason, transitioning protocols do not allow for an overlap between cangrelor and clopidogrel, and requires loading of clopidogrel simultaneously with cangrelor infusion cessation.

However, it is pertinent to note that the time to peak action for clopidogrel of 2-6 hours, while cangrelor has a rapid elimination time of nearly 30 minutes from infusion cessation. This will inevitably introduce a short period of partial platelet reactivation during transition from cangrelor to clopidogrel. Other antiplatelets that do not target the P2Y12 receptor, like prasugrel and ticagrelor, can be loaded 1 to 2 hours before the cessation of cangrelor infusion, eliminating the possibility of partial platelet reactivation. In our initial experience with cangrelor, clopidogrel was our default transition agent. One patient in the cangrelor group who had an intracranial stent placed and was transitioned to clopidogrel had a postoperative course complicated by in-stent thrombosis within 24 hours post-procedure, which led to poor outcome with the patient’s eventual death. The transient period of partial platelet reactivity during the transition from cangrelor to clopidogrel likely increases the risk of intracranial stent occlusion, which carry a higher risk of thrombotic occlusion compared to extracranial stents.^38^ Subsequently, we modified our institutional practice such that ticagrelor is now the transition agent of choice, after intracranial stenting.

A subset of patients in both the eptifibatide and cangrelor groups did not receive any transition agent after administration of the medication. This was primarily in patients for whom no stent was placed and for whom the antiplatelet agent was used as an adjuvant for distal recanalization. Additionally, some patients were not continued on antiplatelet treatment because of the occurrence of hemorrhagic complications or a change in the overall goals of care of treatment. In part due to these considerations, significant univariate differences were noted in the choice of transition agent between the two groups. All our patients received aspirin, unless hemorrhagic conversion occurred.

The use of cangrelor does raise potential financial concerns. One 50mg vial of cangrelor costs approximately $774 at our institution.^39^ Eptifibatide is notably cheaper, as 75mg bolus vials cost $0.61 per vial when purchased in bulk and 20mg infusion vials cost $1.88. To minimize such costs, patients are often rapidly transitioned to an oral transition agent with use of a nasogastric tube if the patient continues to be intubated or is unable to swallow. The therapeutic benefits of early antiplatelet administration even by nasogastric tube are well documented in the literature through large randomized trials such as CAST, but minimizing the duration of cangrelor use also reduces overall cost of administration.^40,41^ Additionally, the cost savings from reduced use of ancillary services during hospitalization and improved functional outcomes in long-term follow-up have yet to be elucidated and may offset the cost of utilizing cangrelor.^42^ From a cardiological perspective, one 2022 study at a high-volume center demonstrated a 12.8% reduction in cost as usage of cangrelor increased from 11% to 32%, primarily driven by reductions in complications.^43^ This cost analysis in the neurointerventional setting is currently under study at our institution.

There are some limitations to the study. As this was a retrospective analysis, the patients were not randomized to receive cangrelor or eptifibatide and selection bias may be present. The number of patients who received cangrelor was relatively small compared to the number who received eptifibatide. We attempted to control for any biases or confounding variables with multivariate analyses. Some patients were lost to follow-up and could not be contacted in a timely fashion to obtain a 90-day mRS. Lastly, not all patients received 24-hour vascular imaging confirming vessel or stent patency during the same hospitalization as their EVT, and so our ability to make accurate conclusions about the rate of thrombotic complications was limited.

## Conclusion

In patients receiving endovascular thrombectomy for management of acute ischemic stroke, cangrelor use was associated with a significantly reduced risk of hemorrhagic complications and symptomatic ICH without an increase in risk of thrombotic complications. The use of cangrelor was associated with more favorable functional outcomes for patients during hospitalization, at discharge and at 90 days when compared to GPI use. Future randomized and prospective studies are warranted.

## Data Availability

The corresponding (AD) and senior author (HS) had full access to all data collected and analyzed in the study, and take responsibility for its analysis and integrity. Data may be made available at reasonable request.

## Non-standard Abbreviations and Acronyms

AOR: adjusted odds ratio
ASPECTS: Alberta Stroke Program Early CT Score
CAD: coronary artery disease
CAST: Chinese Acute Stroke Trial
CHF: congestive heart failure
CI: confidence interval
CT: computerized tomography
ECASS: European Cooperative Acute Stroke Study
EVT: endovascular thrombectomy
GPI: glycoprotein IIb-IIIa protein inhibitor
HT: hemorrhagic transformation
ICAD: intracranial atherosclerotic disease
ICU: intensive care unit
IQR: interquartile range
IRB: Institutional Review Board
IV: intravenous
MRI: magnetic resonance imaging
mRS: modified Rankin Scale
NIHSS: National Institutes of Health Stroke Scale
OR: odds ratio
PH: parenchymal hematoma
PRU: platelet reactivity unit
VIF: variance inflation factor

## References

1. Powers WJ, Rabinstein AA, Ackerson T, et al. Guidelines for the Early Management of Patients With Acute Ischemic Stroke: 2019 Update to the 2018 Guidelines for the Early Management of Acute Ischemic Stroke: A Guideline for Healthcare Professionals From the American Heart Association/American Stroke Association. Stroke. 2019;50(12):e344–e418. doi:10.1161/STR.0000000000000211

2. Nogueira RG, Jadhav AP, Haussen DC, et al. Thrombectomy 6 to 24 Hours after Stroke with a Mismatch between Deficit and Infarct. N Engl J Med. 2018;378(1):11–21. doi:10.1056/NEJMoa1706442

3. Albers GW, Marks MP, Kemp S, et al. Thrombectomy for Stroke at 6 to 16 Hours with Selection by Perfusion Imaging. N Engl J Med. 2018;378(8):708–718. doi:10.1056/NEJMoa1713973

4. Goyal M, Menon BK, van Zwam WH, et al. Endovascular thrombectomy after large-vessel ischaemic stroke: a meta-analysis of individual patient data from five randomised trials. Lancet Lond Engl. 2016;387(10029):1723–1731. doi:10.1016/S0140-6736(16)00163-X

5. Oliveira R, Correia MA, Marto JP, et al. Reocclusion after successful endovascular treatment in acute ischemic stroke: systematic review and meta-analysis. J Neurointerventional Surg. Published online November 3, 2022:jnis-2022-019382. doi:10.1136/jnis-2022-019382

6. Mazighi M, Labreuche J, Gongora-Rivera F, Duyckaerts C, Hauw JJ, Amarenco P. Autopsy Prevalence of Intracranial Atherosclerosis in Patients With Fatal Stroke. Stroke. 2008;39(4):1142–1147. doi:10.1161/STROKEAHA.107.496513

7. Hallevi H, Chernyshev OY, Khoury RE, et al. Intracranial Atherosclerosis Is Associated with Progression of Neurological Deficit in Subcortical Stroke. Cerebrovasc Dis. 2012;33(1):64–68. doi:10.1159/000333388

8. Eker OF, Bühlmann M, Dargazanli C, et al. Endovascular Treatment of Atherosclerotic Tandem Occlusions in Anterior Circulation Stroke: Technical Aspects and Complications Compared to Isolated Intracranial Occlusions. Front Neurol. 2018;9. Accessed April 17, 2023. https://www.frontiersin.org/articles/10.3389/fneur.2018.01046

9. Kang DH, Yoon W, Kim SK, et al. Endovascular treatment for emergent large vessel occlusion due to severe intracranial atherosclerotic stenosis. J Neurosurg. Published online June 1, 2018:1–8. doi:10.3171/2018.1.JNS172350

10. Vaduganathan M, Harrington RA, Stone GW, et al. Evaluation of Ischemic and Bleeding Risks Associated With 2 Parenteral Antiplatelet Strategies Comparing Cangrelor With Glycoprotein IIb/IIIa Inhibitors: An Exploratory Analysis From the CHAMPION Trials. JAMA Cardiol. 2017;2(2):127–135. doi:10.1001/jamacardio.2016.4556

11. Walsh RD, Barrett KM, Aguilar MI, et al. Intracranial hemorrhage following neuroendovascular procedures with abciximab is associated with high mortality: a multicenter series. Neurocrit Care. 2011;15(1):85–95. doi:10.1007/s12028-010-9338-1

12. De Luca L, Steg PG, Bhatt DL, Capodanno D, Angiolillo DJ. Cangrelor: Clinical Data, Contemporary Use, and Future Perspectives. J Am Heart Assoc. 2021;10(13):e022125. doi:10.1161/JAHA.121.022125

13. Harrington RA, Stone GW, McNulty S, et al. Platelet Inhibition with Cangrelor in Patients Undergoing PCI. N Engl J Med. 2009;361(24):2318–2329. doi:10.1056/NEJMoa0908628

14. Bhatt DL, Stone GW, Mahaffey KW, et al. Effect of platelet inhibition with cangrelor during PCI on ischemic events. N Engl J Med. 2013;368(14):1303–1313. doi:10.1056/NEJMoa1300815

15. Bhatt DL, Lincoff AM, Gibson CM, et al. Intravenous Platelet Blockade with Cangrelor during PCI. N Engl J Med. 2009;361(24):2330–2341. doi:10.1056/NEJMoa0908629

16. Giossi A, Pezzini A, Del Zotto E, et al. Advances in antiplatelet therapy for stroke prevention: the new P2Y12 antagonists. Curr Drug Targets. 2010;11(3):380–391. doi:10.2174/138945010790711987

17. Linfante I, Ravipati K, Starosciak AK, Reyes D, Dabus G. Intravenous cangrelor and oral ticagrelor as an alternative to clopidogrel in acute intervention. J Neurointerventional Surg. 2021;13(1):30–32. doi:10.1136/neurintsurg-2020-015841

18. Elhorany M, Lenck S, Degos V, et al. Cangrelor and Stenting in Acute Ischemic Stroke : Monocentric Case Series. Clin Neuroradiol. 2021;31(2):439–448. doi:10.1007/s00062-020-00907-0

19. Aguilar-Salinas P, Agnoletto GJ, Brasiliense LBC, et al. Safety and efficacy of cangrelor in acute stenting for the treatment of cerebrovascular pathology: preliminary experience in a single-center pilot study. J Neurointerventional Surg. 2019;11(4):347–351. doi:10.1136/neurintsurg-2018-014396

20. Marnat G, Finistis S, Delvoye F, et al. Safety and Efficacy of Cangrelor in Acute Stroke Treated with Mechanical Thrombectomy: Endovascular Treatment of Ischemic Stroke Registry and Meta-analysis. AJNR Am J Neuroradiol. 2022;43(3):410–415. doi:10.3174/ajnr.A7430

21. Salahuddin H, Dawod G, Zaidi SF, Shawver J, Burgess R, Jumaa MA. Safety of Low Dose Intravenous Cangrelor in Acute Ischemic Stroke: A Case Series. Front Neurol. 2021;12:636682. doi:10.3389/fneur.2021.636682

22. Barber PA, Demchuk AM, Zhang J, Buchan AM. Validity and reliability of a quantitative computed tomography score in predicting outcome of hyperacute stroke before thrombolytic therapy. ASPECTS Study Group. Alberta Stroke Programme Early CT Score. Lancet Lond Engl. 2000;355(9216):1670–1674. doi:10.1016/s0140-6736(00)02237-6

23. Hacke W, Kaste M, Fieschi C, et al. Intravenous thrombolysis with recombinant tissue plasminogen activator for acute hemispheric stroke. The European Cooperative Acute Stroke Study (ECASS). JAMA. 1995;274(13):1017–1025.

24. Hacke W, Kaste M, Fieschi C, et al. Randomised double-blind placebo-controlled trial of thrombolytic therapy with intravenous alteplase in acute ischaemic stroke (ECASS II). Second European-Australasian Acute Stroke Study Investigators. Lancet Lond Engl. 1998;352(9136):1245–1251. doi:10.1016/s0140-6736(98)08020-9

25. von Kummer R, Broderick JP, Campbell BCV, et al. The Heidelberg Bleeding Classification. Stroke. 2015;46(10):2981–2986. doi:10.1161/STROKEAHA.115.010049

26. Calculating & Preparing a Dose. KENGREAL® (cangrelor). Accessed September 15, 2023. https://kengreal.com/dosing-and-administration/

27. Cervo A, Ferrari F, Barchetti G, et al. Use of Cangrelor in Cervical and Intracranial Stenting for the Treatment of Acute Ischemic Stroke: A “Real Life” Single-Center Experience. AJNR Am J Neuroradiol. 2020;41(11):2094–2099. doi:10.3174/ajnr.A6785

28. Lipinski MJ, Lee RC, Gaglia MA, et al. Comparison of heparin, bivalirudin, and different glycoprotein IIb/IIIa inhibitor regimens for anticoagulation during percutaneous coronary intervention: A network meta-analysis. Cardiovasc Revascularization Med Mol Interv. 2016;17(8):535–545. doi:10.1016/j.carrev.2016.09.011

29. Marnat G, Delvoye F, Finitsis S, et al. A Multicenter Preliminary Study of Cangrelor following Thrombectomy Failure for Refractory Proximal Intracranial Occlusions. AJNR Am J Neuroradiol. 2021;42(8):1452–1457. doi:10.3174/ajnr.A7180

30. Delvoye F, Maier B, Escalard S, et al. Antiplatelet Therapy During Emergent Extracranial Internal Carotid Artery Stenting: Comparison of Three Intravenous Antiplatelet Perioperative Strategies. J Stroke Cerebrovasc Dis Off J Natl Stroke Assoc. 2021;30(2):105521. doi:10.1016/j.jstrokecerebrovasdis.2020.105521

31. Cagnazzo F, Radu RA, Derraz I, et al. Efficacy and safety of low dose intravenous cangrelor in a consecutive cohort of patients undergoing neuroendovascular procedures. J Neurointerventional Surg. Published online March 15, 2023:jnis-2023-020094. doi:10.1136/jnis-2023-020094

32. Abdennour L, Sourour N, Drir M, et al. Preliminary Experience with Cangrelor for Endovascular Treatment of Challenging Intracranial Aneurysms. Clin Neuroradiol. 2020;30(3):453–461. doi:10.1007/s00062-019-00811-2

33. Cheddad El Aouni M, Magro E, Abdelrady M, Nonent M, Gentric JC, Ognard J. Safety and Efficacy of Cangrelor Among Three Antiplatelet Regimens During Stent-Assisted Endovascular Treatment of Unruptured Intracranial Aneurysm: A Single-Center Retrospective Study. Front Neurol. 2022;13:727026. doi:10.3389/fneur.2022.727026

34. Cagnazzo F, Cappucci M, Dargazanli C, et al. Treatment of Distal Anterior Cerebral Artery Aneurysms with Flow-Diverter Stents: A Single-Center Experience. AJNR Am J Neuroradiol. 2018;39(6):1100–1106. doi:10.3174/ajnr.A5615

35. Entezami P, Holden DN, Boulos AS, et al. Cangrelor dose titration using platelet function testing during cerebrovascular stent placement. Interv Neuroradiol J Peritherapeutic Neuroradiol Surg Proced Relat Neurosci. 2021;27(1):88–98. doi:10.1177/1591019920936923

36. Michelson AD. P2Y12 Antagonism. Arterioscler Thromb Vasc Biol. 2008;28(3):s33–s38. doi:10.1161/ATVBAHA.107.160689

37. Müller I, Besta F, Schulz C, Massberg S, Schönig A, Gawaz M. Prevalence of clopidogrel non-responders among patients with stable angina pectoris scheduled for elective coronary stent placement. Thromb Haemost. 2003;89(5):783–787.

38. Riedel CH, Tietke M, Alfke K, Stingele R, Jansen O. Subacute Stent Thrombosis in Intracranial Stenting. Stroke. 2009;40(4):1310–1314. doi:10.1161/STROKEAHA.108.531400

39. AnalySource - Premier Drug Pricing Services. Accessed September 15, 2023. https://www.analysource.com/

40. Chen ZM. CAST: randomised placebo-controlled trial of early aspirin use in 20 000 patients with acute ischaemic stroke. The Lancet. 1997;349(9066):1641–1649. doi:10.1016/S0140-6736(97)04010-5

41. Minhas JS, Chithiramohan T, Wang X, et al. Oral antiplatelet therapy for acute ischaemic stroke. Cochrane Database Syst Rev. 2022;2022(1):CD000029. doi:10.1002/14651858.CD000029.pub4

42. Yun AN, Toyoda AY, Solomon EJ, Roberts RJ, Ji CS. Safety and Efficacy of Periprocedural Bridging With Cangrelor Versus Eptifibatide. J Cardiovasc Pharmacol. 2022;79(3):383–389. doi:10.1097/FJC.0000000000001192

43. Jensen IS, Wu E, Cyr PL, et al. Cost-Consequence Analysis of Using Cangrelor in High Angiographic Risk Percutaneous Coronary Intervention Patients: A US Hospital Perspective. Am J Cardiovasc Drugs. 2022;22(1):93–104. doi:10.1007/s40256-021-00491-9

